# Randomized attention-placebo controlled trial of a digital self-management platform for adult asthma

**DOI:** 10.1101/2023.07.19.23292865

**Authors:** Aaron Kandola, Kyra Edwards, Joris Straatman, Bettina Dührkoop, Bettina Hein, Joseph F Hayes

**Affiliations:** MRC Unit of Lifelong Health and Aging, UCL; juli Health; Division of Psychiatry, UCL; Camden and Islington NHS Foundation Trust

**Keywords:** asthma, mobile health, self-management, randomized controlled trial

## Abstract

**Background:** Asthma is one of the most common chronic conditions worldwide, with a substantial individual, societal and healthcare burden. Digital apps hold promise as a highly accessible, low-cost method of enhancing self-management in asthma, which is critical to effective asthma control.

**Objective:** We conducted a fully remote trial to assess the efficacy of juli, a commercially available smartphone self-management platform for asthma.

**Methods:** We conducted a pragmatic single-blind, randomized controlled trial of juli for asthma management. Our study included participants aged 18 and above with asthma and had an asthma control test (ACT) score of 19 or less (indicating poorly controlled asthma) at the beginning of the trial. Participants were randomized (1:1 ratio) to receive juli for eight weeks or a limited attention-placebo control version of the app. The primary endpoint was the difference in ACT scores after eight weeks. Secondary endpoints included eight-week remission (ACT score greater than 19), minimal clinically important difference (an improvement of three or more points on the ACT), worsening of asthma, and health-related quality of life. The primary analysis included participants using the app for eight weeks (per-protocol), a secondary analysis used modified intention-to-treat.

**Results:** We randomized 411 participants between May 2021 and April 2023: 152 engaged with the app for eight weeks and were included in the per-protocol analysis, 262 completed the week two outcome assessment and were included in the modified intention-to-treat analysis.

In the per-protocol analysis, the intervention group had a higher mean ACT score (17.93, standard deviation = 4.72) than the control group (16.24, standard deviation = 5.78) by week eight (baseline adjusted β-coefficient 1.91, 95% confidence intervals = 0.31 to 3.51, p=0.020). Participants using juli had greater odds of achieving or exceeding the minimal clinically important difference at eight weeks (adjusted odds ratio = 2.38, 95% confidence intervals = 1.20 to 4.70, p=0.013). There were no between-group differences in the other secondary outcomes at eight weeks. The results from the modified intention-to-treat analysis were similar.

**Conclusions:** Users of juli had improved asthma symptom control over eight weeks compared with users of a version of the app with limited functionality. These findings suggest that juli is an effective digital self-management platform that could augment existing care pathways for asthma.

**Trial Registration:** The trial was pre-registered (ISRCTN87679686).

## Introduction

Asthma is one of the most common chronic conditions worldwide, with an increasing prevalence, currently affecting one in ten people at some time during their lives.^1,2^ The inflammatory disease causes mild-to-severe respiratory symptoms, including shortness of breath, chest tightness, wheezing, and cough. It significantly burdens patients and healthcare services, including long-term treatment, emergency care, and hospitalizations that will cost the United States economy an estimated $300 billion over the next 20 years in direct healthcare expenditure.^3^ Effective asthma control is necessary to reduce these costs and improve the quality of life in people with the condition.

Asthma is currently not curable, so its management is based on achieving symptom control and reducing the frequency and severity of exacerbations.^4^ This involves the use of inhaled anti-inflammatory medications and the avoidance of asthma triggers. Symptom control is associated with improved quality of life, reduced healthcare costs and better work performance.^5^ However, a significant proportion of individuals with asthma have suboptimal control because of poor adherence to medication, insufficient recognition of triggers, comorbidities (such as rhinitis or obesity), health behaviors (such as smoking) and inadequate information about treatment.^6^ Digital interventions may address some of these treatment challenges by enabling people with asthma to more easily and consistently self-manage their condition compared to existing treatment plans. For example, digital interventions can offer timely reminders to improve medication adherence or real-time feedback to identify and adapt to possible triggers and health behaviors.

A 2017 systematic review and meta-analysis of randomized controlled trials (RCTs) of digital interventions in adults with asthma found modest improvements in asthma control overall, but results were inconsistent.^7^ A similar 2018 review of RCTs and observational studies concluded that apps were more effective than other types of digital interventions, such as web-based interventions.^8^ Studies published since these reviews have generally been feasibility trials or small underpowered RCTs.^9-11^ A 2022 Cochrane review examined the effect of digital interventions for asthma medication adherence, concluding they were likely to be useful in poorly adherent populations.^12,13^ However, the review did not examine other components of digital self-management like symptom tracking, which are included in many commercially available apps. These commercially available apps are widespread but lack the rigorous assessment appropriate for clinical interventions. Many of the research-grade apps included in these reviews are not commercially available to patients. Other challenges include low levels of retention and engagement with digital health apps in trials and in real-world use.^14^

We aimed to address the fundamental issue that commercially available apps are widespread but require sufficient evaluation of their effectiveness by conducting an RCT of juli. This is a digital health app that aims to support people with asthma by combining numerous approaches that have been shown effective in research-grade apps for asthma, including symptom tracking, medication reminders, trigger identification (including geolocated weather, pollen and air pollution data), data visualization of respiratory symptoms, mood, exercise, activity, sleep, and heart rate variability and behavioral activation recommendations about how to improve these parameters.^15,16^ Our RCT was fully remote, increasing time-efficiency, cost-effectiveness, and reach. We hypothesized that participants who were randomized to receive juli would have a greater reduction in asthma symptoms at eight weeks than those randomized to attention-placebo control.

## Methods

### Study design and participants

We conducted a fully remote pragmatic single-blind, placebo controlled randomized controlled trial to test the efficacy of juli in people with asthma. The trial was open to individuals from anywhere in the world providing they were aged 18 to 65, English-speaking, had access to a smartphone, and a diagnosis of asthma. We also only included people with asthma symptoms that were moderately or poorly controlled based upon a score of 19 or lower on the asthma control test (ACT) at baseline. Recruitment ran from May 2021 until April 2023 and included self-help groups for asthma, online advertisements and social media posts. All participants supplied written informed consent within the app. The University College London Ethics Committee gave full ethical approval (ID: 19413/001). The trial was entered on the ISRCTN registry (ISRCTN87679686). At the same time we were running an RCT of the juli app for depression. This RCT had a similar design and analysis.^17^

### Randomization and masking

We assigned participants in a 1:1 ratio to either an attention-placebo control or the full version of juli. We automated and conducted randomization within the app, employing random block sizes ranging from four to eight. To ensure data integrity, the treatment allocation was concealed from both the research team and independent statisticians until the analysis was finalized.

### Procedures

The juli app was developed by gamification experts in collaboration with patients, a psychiatrist, and a pulmonologist. Our trial used a full version of the juli app for the intervention group and a limited version in the attention-placebo control group. Participants with the complete juli app received automatic prompts to open the app each day at a user-inputted time. The app asked participants about how their asthma was affecting them on a five face emoji scale, their emergency inhaler usage that day, how often they had a shortness of breath episode and whether they woke in the night due to shortness of breath. Individuals could also track various factors they regarded as relevant to their asthma symptoms, such as tobacco smoke exposure.^16^ The app connects to smart peak flow meters or participants can enter this information manually.

The app presented participants with regular, geolocated weather, pollen and air pollution data relevant to their asthma.^18^ Users could also access passively gathered smartphone and wearable sensor information on relevant health-related factors, such as workouts, activity, heart rate variability, menstrual cycle, and sleep. Users could check this information daily and understand associations with their asthma.^19-21^ The app also uses behavioral activation techniques to provide personalized recommendations about these factors to encourage healthy behaviors. The app includes customizable medication reminders to improve medication adherence.^22^ The juli app also encourages participants to use the positive affect journaling function.^23^ The design of the juli app guides participants towards all elements of the app but allows them the flexibility to choose where they want to engage.

Participants in the control arm had a limited version of the app. The app prompted participants to open it each day and rate how they were feeling on the five face emoji scale, but they did not have access to any further functionality or intervention. There was no change to usual care in either arm.

Participants in both arms completed baseline assessments and follow-up assessments at two, four, six and eight weeks remotely from within the app. Assessments included the ACT for asthma symptoms and the 12-Item Short Form Health Survey (SF-12) for health-related quality of life. The ACT is a widely used, clinically validated, self-completed asthma symptom scale that is responsive to change with scores ranging from 5-25.^24^ A cut-off score of 19 or less identifies patients with poorly controlled asthma. The SF-12 is a self-reported measure assessing the impact of health on an individual’s everyday life. Scores range from 0-100 with higher scores indicating better quality-of-life.^25^

### Outcomes

The total ACT score at eight weeks was our primary endpoint. Secondary endpoints were: continuous ACT score at two, four, six, and eight weeks in a repeated measures analysis using mixed-effect models; remission, defined as a score of >19 at eight weeks; remission at two, four, six, and eight weeks in a repeated measures analysis; SF-12 physical and mental component scores at eight weeks and; SF-12 physical and mental component scores at four and eight weeks in a repeated measures analysis.

We added achieving a minimal clinically important difference (MCID) at eight weeks (3-point increase on the ACT),^26^ and a worsening of asthma symptoms (i.e., a decrease in ACT scores from baseline) as post-hoc outcomes.

### Sample Size estimation

The best MCID estimate for the ACT is between 2.2 and 3.0 (standard deviation (SD) = 3.1 to 4.7)^26^. A two-sided 5% significance level at 80% power requires a total sample size of 146 for an MCID of 3. We aimed to recruit 90 participants per arm, allowing for 23% attrition.^27^

### Statistical analyses

We pre-printed the analysis plan on UCL Discovery (https://discovery.ucl.ac.uk/id/eprint/10129351/) and pre-registered the RCT on the ISRCTN registry with a description of the primary and secondary outcomes before the trial started. In reporting and analyzing our data we followed the Consolidated Standards of Reporting Trials (CONSORT) guidelines.^28^

Our primary endpoint was the difference in total ACT score at eight weeks between the control and intervention groups in a per-protocol analysis. We estimated this difference with a linear regression model adjusted for baseline ACT and any imbalanced baseline covariates. We used logistic regression to calculate the odds ratio (OR) of remission at eight weeks (ACT>19), achieving MCID (≥3 point ACT improvement) and worsening of asthma, adjusting for baseline ACT. We completed the repeat measures analyses using linear or logistic mixed-effect models adjusting for ACT at baseline.

We repeated analysis of all endpoints in a modified intention-to-treat analysis. This analysis included all randomized participants with a complete baseline and week two ACT score, dropping participants who were randomized but never used the app (Figure 1). We imputed the missing ACT scores first using multiple imputation models and then using the last observation carried forward.^29^ The multiple imputation models included predictive mean matching with five nearest neighbors and 50 iterations. This method means that only plausible values are imputed, and is more robust to model misspecification than fully parametric imputation.^30^

**Figure 1.**
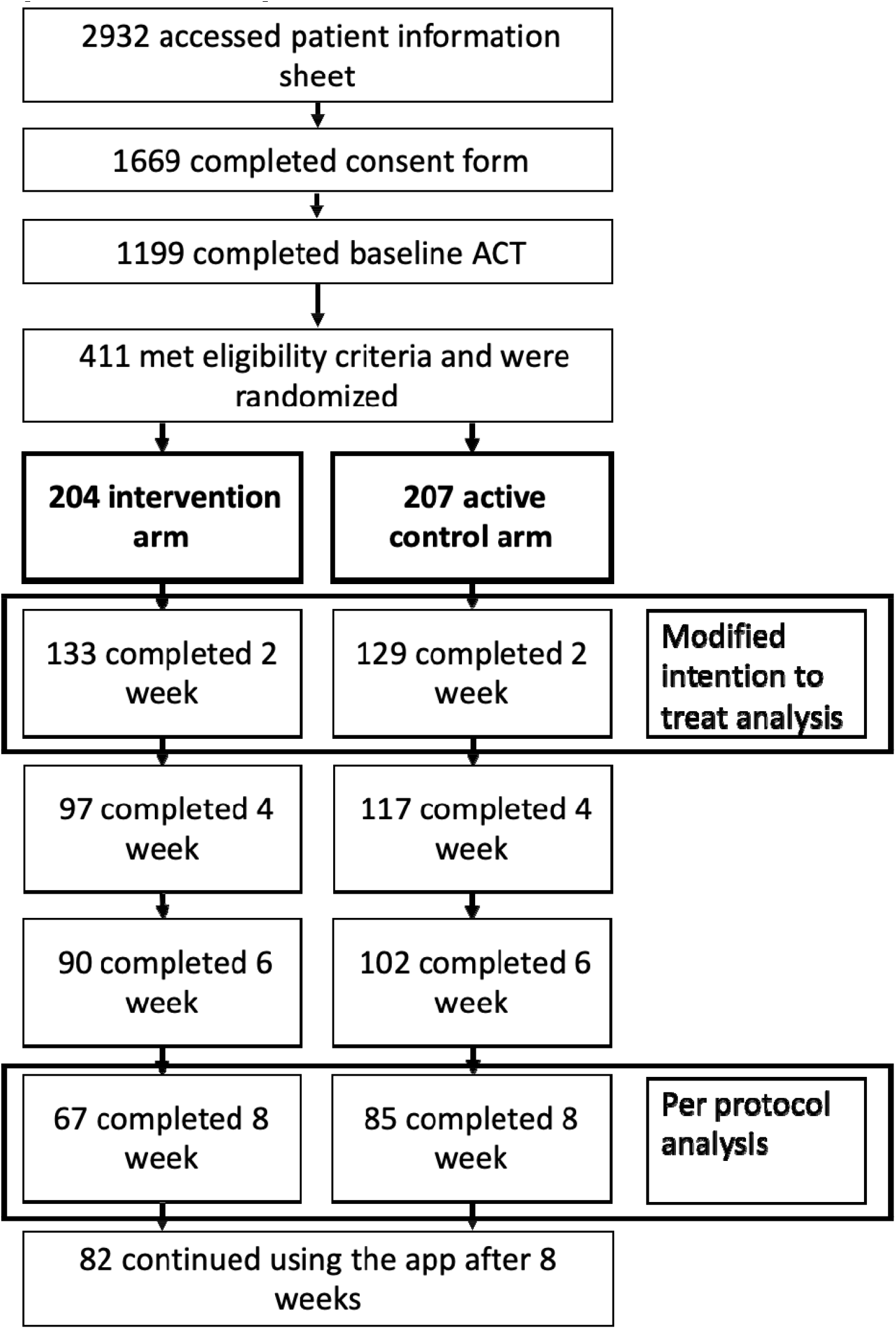
CONSORT diagram.

An independent statistician (KE) with no conflicts of interest with the company providing juli completed the analyses. All analyses we conducted using Stata (Version 17) and R (Version 4.3.1 for Windows).

## Results

### Per-protocol analysis

We recruited 152 participants who remained in the trial for eight weeks and contributed to our per-protocol analysis (Figure 1). Participants were mostly females who had been diagnosed with by a physician more than five years ago and had ongoing contact with a doctor about their asthma (Table 1). Participants had a mean baseline ACT score of 12.84 (SD = 4.00).

**Table 1.**
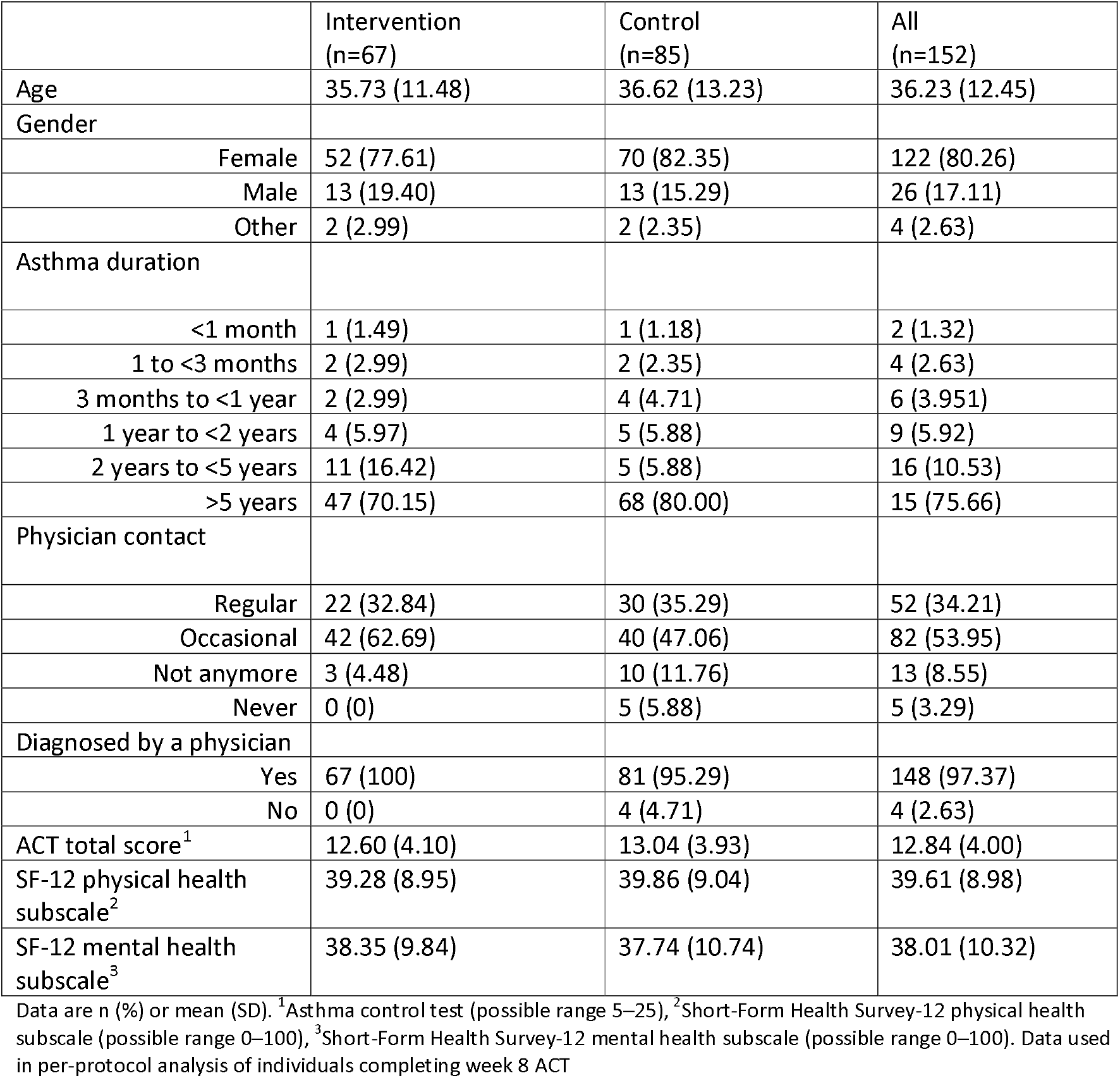
Baseline characteristics.

|  | Intervention<br>(n=67) | Control<br>(n=85) | All<br>(n=152) |
| --- | --- | --- | --- |
| Age | 35.73 (11.48) | 36.62 (13.23) | 36.23 (12.45) |
| Gender |  |  |  |
| Female | 52 (77.61) | 70 (82.35) | 122 (80.26) |
| Male | 13 (19.40) | 13 (15.29) | 26 (17.11) |
| Other | 2 (2.99) | 2 (2.35) | 4 (2.63) |
| Asthma duration |  |  |  |
| <1 month | 1 (1.49) | 1 (1.18) | 2 (1.32) |
| 1 to <3 months | 2 (2.99) | 2 (2.35) | 4 (2.63) |
| 3 months to <1 year | 2 (2.99) | 4 (4.71) | 6 (3.951) |
| 1 year to <2 years | 4 (5.97) | 5 (5.88) | 9 (5.92) |
| 2 years to <5 years | 11 (16.42) | 5 (5.88) | 16 (10.53) |
| >5 years | 47 (70.15) | 68 (80.00) | 15 (75.66) |
| Physician contact |  |  |  |
| Regular | 22 (32.84) | 30 (35.29) | 52 (34.21) |
| Occasional | 42 (62.69) | 40 (47.06) | 82 (53.95) |
| Not anymore | 3 (4.48) | 10 (11.76) | 13 (8.55) |
| Never | 0 (0) | 5 (5.88) | 5 (3.29) |
| Diagnosed by a physician |  |  |  |
| Yes | 67 (100) | 81 (95.29) | 148 (97.37) |
| No | 0 (0) | 4 (4.71) | 4 (2.63) |
| ACT total score<sup>1</sup> | 12.60 (4.10) | 13.04 (3.93) | 12.84 (4.00) |
| SF-12 physical health subscale<sup>2</sup> | 39.28 (8.95) | 39.86 (9.04) | 39.61 (8.98) |
| SF-12 mental health subscale<sup>3</sup> | 38.35 (9.84) | 37.74 (10.74) | 38.01 (10.32) |

Data are n (%) or mean (SD). <sup>1</sup>Asthma control test (possible range 5–25), <sup>2</sup>Short-Form Health Survey-12 physical health subscale (possible range 0–100), <sup>3</sup>Short-Form Health Survey-12 mental health subscale (possible range 0–100). Data used in per-protocol analysis of individuals completing week 8 ACT

After eight weeks, the intervention group participants had a mean ACT score of 17.93 (SD = 4.72) compared with 16.24 (SD = 5.78) in the control group (Figure 2). After adjusting for baseline ACT score, the intervention group showed a greater improvement in symptom scores at eight weeks than those in the control group (adjusted β-coefficient = 1.91, 95% confidence interval (CI) = 0.31 to 3.51, p=0.020). After adjusting for additional imbalanced baseline characteristics (physician contact), the improvement was 2.01 points on the ACT (95%CI = 0.48 to 3.53, p=0.010).

**Figure 2.**
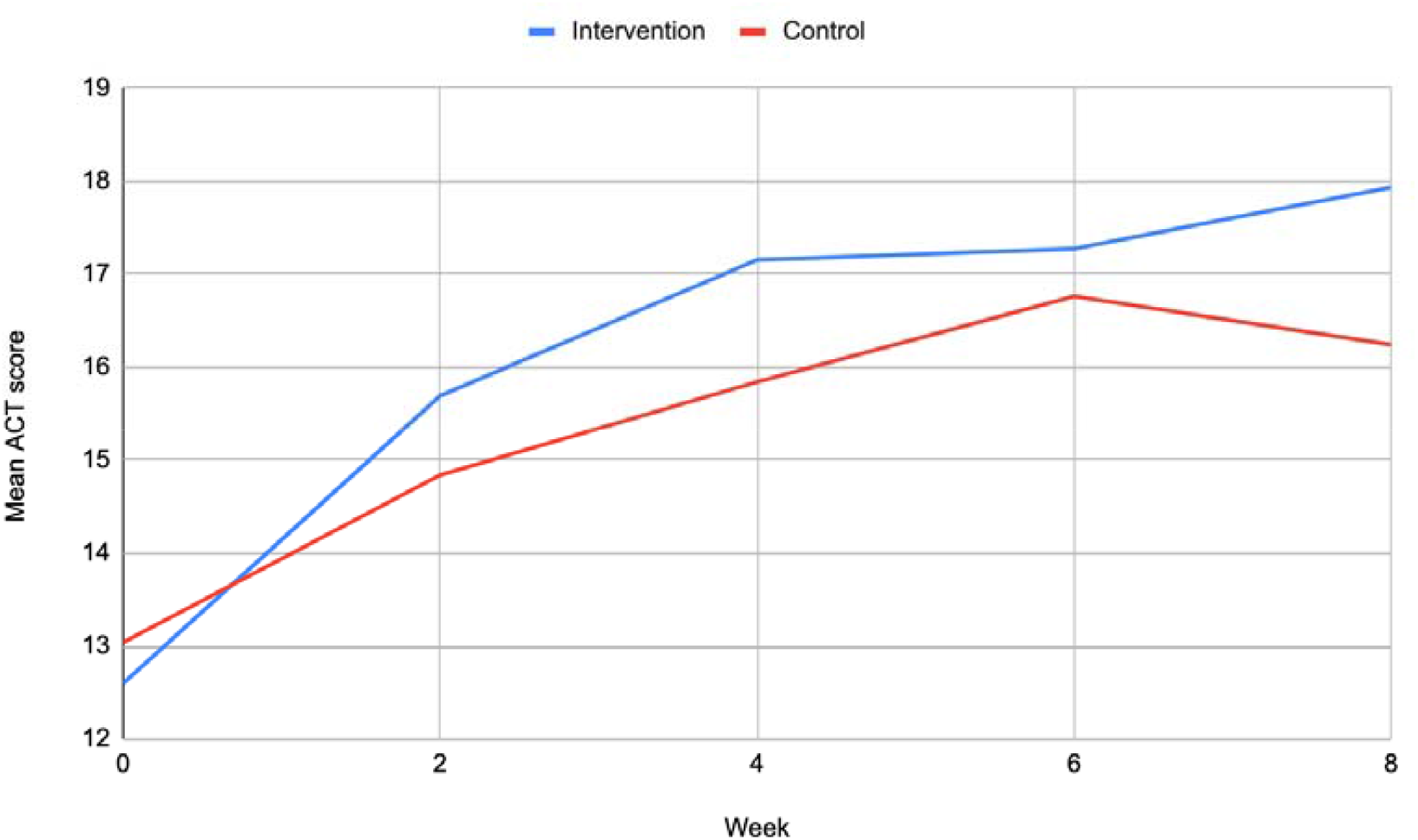
Mean change in ACT score over 8 weeks.

The chance of being in remission by week eight did not differ between the intervention and control group, after accounting for baseline asthma severity. However, participants in the intervention group were more likely to experience a MCID (adjusted OR = 2.38, 95%CI = 1.20 to 4.70, p=0.013) than those in the control group. This effect was consistent across the two, four, six, and eight-week assessments (Supplement Table 2). The odds of worsening symptoms were similar in both arms (adjusted OR = 0.55, 95%CI = 0.23 to 1.32, p=0.182). There were no between group differences in SF-12 mental or physical component scores.

### Intention-to-treat analysis

Of those randomized, 133 participants in the intervention group and 129 in the control group completed the baseline and week two ACT (Supplement Table 1), and were included in the modified intention-to-treat analysis. The baseline characteristics of participants in the intervention and control groups were similar to the per-protocol analysis. Following multiple imputation of missing outcomes, there was a greater improvement in ACT scores in the intervention group than in the active control group (adjusted β-coefficient = 1.56, 95%CI = 0.32 to 2.79, p=0.013) (Supplement Table 2). MCID was more common in the intervention group than the control group (adjusted OR = 2.17, 95%CI = 1.25 to 3.78, p=0.006). Both arms had similar odds of remission, worsening of symptoms, and SF-12 scores. Results from the last observation carried forward data set were consistent with the per-protocol and multiply imputed results.

## Discussion

### Principal results

Our primary analysis showed that juli users had a greater improvement in asthma symptoms at eight weeks compared to an attention-placebo control. The mean improvement in the intervention group was 5.33 compared with 3.20 in the control group. This total improvement and the difference between arms is consistent with a clinically important effect of juli on asthma control.26 Participants assigned to juli had more than twice the odds of a 3-point (MCID) or greater improvement on the ACT. The results from our multilevel models covering outcomes from two to eight weeks and the modified intention-to-treat analysis with all individuals who were randomized and used the app for at least two weeks were consistent with these primary findings.

Participants entering our trial were chronically unwell with potentially difficult-to-treat asthma. For example, participants had a mean baseline ACT score indicating very poorly controlled asthma, and most reported having asthma for several years with routine physician contact. The results of this trial indicate that juli can augment the treatment of poorly-controlled asthma as indicated by improved ACT scores over eight weeks. There is consistent evidence that low ACT scores are associated with rescue medication use, asthma exacerbations, reduced lung function, reduced asthma-specific quality of life, sleep, work and productivity.^31^ Increases in ACT scores are associated with decreased healthcare utilization and healthcare costs.^31^

It is unclear which component of juli resulted in improved ACT scores, but participants likely chose elements that suited them, which is a strength of juli’s design. Meta-analyses results support the self-management of asthma with digital approaches. However, interventions included in reviews are often not available to patients, typically because they are unavailable commercially or through healthcare providers. A combination of evidence-based approaches to support asthma self-management are used by juli and it is available in Android and Apple formats globally. It is a highly accessible platform for people with asthma, and our trial provides methodologically robust evidence of its efficacy in managing asthma. Additional research is required to understand the most cost-effective support procedures to improve adherence to digital self-management tools and how best to integrate them into clinical practice.

### Strengths and limitations

There were several strengths and limitations to this RCT. We successfully and remotely recruited, screened, randomized, treated, and assessed participants worldwide. People could easily participate in the trial as our modified version of the juli app allowed consent, randomization, and assessments to occur within the platform. This facilitated a low-cost global recruitment strategy and a pragmatic trial design with good external validity. However, our focus on reducing participant burden limited the types and richness of data we were able to collect at baseline. For example, we lack relevant information on income, education, and other social determinants of health. Despite this, we did achieve post-randomization balance in recorded characteristics at baseline, indicating successful randomization. Most of the participants were female, reflecting established differences in sex-specific rates of asthma and healthcare utilization in adults.^32^

Participants in our trial had poorly controlled asthma, showing that juli can be effective in these populations. For example, most participants in our study had low ACT scores at baseline, a physician diagnosis, and regular contact with a doctor. However, there is no evidence that participants were no longer accessing traditional care during the trial. Individuals without ongoing care may differentially benefit from digital technologies.

Participants completed the ACT, which is a recommended primary endpoint in clinical trials for asthma interventions.^31^ We also pre-registered our primary and secondary outcome measures along with a full analysis plan, which we adhered to. However, we lacked a broader battery of outcome measures that could have further contextualized our findings and identified possible mechanisms of action.

Attrition was greater than we predicted (63% from randomization to week eight). However, the attrition in our trial follows a similar pattern to other digital RCTs, including for asthma apps, where it mostly occurs between randomization and week two. Dropout rates in previous RCTs have ranged from 20 to 60%.^8^ We continued recruiting until achieving a sufficient number of participants completing the week eight outcome measures. We examined differences in completers versus non-completers (see Table 1 and Supplement Table 1). There were unlikely to be differences between those who dropped out of the study and those who completed it based on their baseline characteristics, including asthma severity. Our modified intention-to-treat and primary analysis findings were similar, suggesting the intervention would have had a similar effect in those who dropped out. The intention-to-treat analysis employed two imputation methods that make different assumptions,^29^ and results were consistent using both methods.

Previous RCTs of digital apps for asthma self-management typically use usual care controls.^7^ This approach can result in a seemingly larger effect size as the control group is less likely to improve and may even experience nocebo effects. We used an active attention-placebo control, potentially reducing the between-group differences observed, but increasing our certainty that the difference observed was due to the effects of the intervention. Participants in our primary analysis used the app for at least eight weeks, which allowed us to isolate the effect of consistent app use on asthma.

## Conclusion

The juli app decreased asthma symptoms within an eight-week period, with an increased chance of participants with poorly controlled asthma achieving an MCID. As such, juli represents a low-risk and low-cost adjunct to the care regimen of individuals with asthma.

## Data Availability

All data produced in the present study are available upon reasonable request to the authors.

## Author Contributions

JFH conceived the study; JFH, AK, BD, BH and JS designed the study; AK, BD and BH collected the data; KE analyzed the data; JFH wrote the initial draft; all authors edited and approved the final manuscript.

## Acknowledgements

AK is supported by the UK Research and Innovation (UKRI) Digital Youth Programme award [MRC project reference MR/W002450/1], which is part of the AHRC/ESRC/MRC Adolescence, Mental Health and the Developing Mind programme. JFH is supported by the UK Research and Innovation grant MR/V023373/1, the University College London Hospitals NIHR Biomedical Research Centre, and the NIHR North Thames Applied Research Collaboration.

## Data Availability

All data produced in the present study are available upon reasonable request to the authors.

## References

1 Nunes, C., Pereira, A. M. & Morais-Almeida, M. Asthma costs and social impact. Asthma research and practice 3, 1–11 (2017).

2 Song, P. et al. Global, regional, and national prevalence of asthma in 2019: a systematic analysis and modelling study. Journal of Global Health 12 (2022).

3 Yaghoubi, M., Adibi, A., Safari, A., FitzGerald, J. M. & Sadatsafavi, M. The projected economic and health burden of uncontrolled asthma in the United States. American journal of respiratory and critical care medicine 200, 1102–1112 (2019).

4 Committee, G. I. f. A. E. in NHLBI/WHO Workshop Report; 2006, GINA.

5 Nguyen, H. V. et al. Association between asthma control and asthma cost: Results from a longitudinal study in a primary care setting. Respirology 22, 454–459 (2017).

6 Braido, F. Failure in asthma control: reasons and consequences. Scientifica 2013 (2013).

7 Hui, C. Y. et al. The use of mobile applications to support self-management for people with asthma: a systematic review of controlled studies to identify features associated with clinical effectiveness and adherence. Journal of the American Medical Informatics Association 24, 619–632 (2017).

8 Unni, E., Gabriel, S. & Ariely, R. A review of the use and effectiveness of digital health technologies in patients with asthma. Annals of Allergy, Asthma & Immunology 121, 680-691. e681 (2018).

9 Ainsworth, B. et al. Feasibility trial of a digital self-management intervention ‘My Breathing Matters’ to improve asthma-related quality of life for UK primary care patients with asthma. BMJ open 9, e032465 (2019).

10 Ljungberg, H. et al. Clinical effect on uncontrolled asthma using a novel digital automated self-management solution: a physician-blinded randomised controlled crossover trial. European Respiratory Journal 54 (2019).

11 Newhouse, N. et al. Randomised feasibility study of a novel experience-based internet intervention to support self-management in chronic asthma. BMJ open 6, e013401 (2016).

12 Chan, A. et al. Digital interventions to improve adherence to maintenance medication in asthma. Cochrane Database of Systematic Reviews (2022).

13 Tinschert, P., Jakob, R., Barata, F., Kramer, J.-N. & Kowatsch, T. The potential of mobile apps for improving asthma self-management: a review of publicly available and well-adopted asthma apps. JMIR mHealth and uHealth 5, e7177 (2017).

14 Camacho-Rivera, M. et al. Evaluating asthma mobile apps to improve asthma self-management: user ratings and sentiment analysis of publicly available apps. JMIR mHealth and uHealth 8, e15076 (2020).

15 Van Lieshout, R. J. & MacQueen, G. Psychological factors in asthma. Allergy, Asthma & Clinical Immunology 4, 1–17 (2008).

16 Gibson, P. G. et al. Self-management education and regular practitioner review for adults with asthma. Cochrane database of systematic reviews 2010 (1996).

17 Kandola, A. et al. Digitally managing depression: a fully remote randomized attention-placebo controlled trial. medRxiv, 2023.2004. 2005.23288184 (2023).

18 Lee, S. W. et al. Short-term effects of multiple outdoor environmental factors on risk of asthma exacerbations: Age-stratified time-series analysis. Journal of Allergy and Clinical Immunology 144, 1542-1550. e1541 (2019).

19 Lehrer, P. M. et al. Heart rate variability biofeedback does not substitute for asthma steroid controller medication. Applied psychophysiology and biofeedback 43, 57–73 (2018).

20 Scheer, F. A. et al. The endogenous circadian system worsens asthma at night independent of sleep and other daily behavioral or environmental cycles. Proceedings of the National Academy of Sciences 118, e2018486118 (2021).

21 Sánchez-Ramos, J. L. et al. Risk factors for premenstrual asthma: a systematic review and meta-analysis. Expert Review of Respiratory Medicine 11, 57–72 (2017).

22 Engelkes, M., Janssens, H. M., de Jongste, J. C., Sturkenboom, M. C. & Verhamme, K. M. Medication adherence and the risk of severe asthma exacerbations: a systematic review. European Respiratory Journal 45, 396–407 (2015).

23 Boggiss, A. L., Consedine, N. S., Brenton-Peters, J. M., Hofman, P. L. & Serlachius, A. S. A systematic review of gratitude interventions: Effects on physical health and health behaviors. Journal of Psychosomatic Research 135, 110165 (2020).

24 Schatz, M. et al. Asthma Control Test: reliability, validity, and responsiveness in patients not previously followed by asthma specialists. Journal of Allergy and Clinical Immunology 117, 549–556 (2006).

25 Jenkinson, C. et al. A shorter form health survey: can the SF-12 replicate results from the SF-36 in longitudinal studies? Journal of Public Health 19, 179–186 (1997).

26 Schatz, M. et al. The minimally important difference of the Asthma Control Test. Journal of Allergy and Clinical Immunology 124, 719-723. e711 (2009).

27 McLean, G. et al. Interactive digital interventions to promote self-management in adults with asthma: systematic review and meta-analysis. BMC pulmonary medicine 16, 1–14 (2016).

28 Schulz, K. F., Altman, D. G. & Moher, D. CONSORT 2010 statement: updated guidelines for reporting parallel group randomised trials. Journal of Pharmacology and pharmacotherapeutics 1, 100–107 (2010).

29 Cro, S., Morris, T. P., Kenward, M. G. & Carpenter, J. R. Sensitivity analysis for clinical trials with missing continuous outcome data using controlled multiple imputation: a practical guide. Statistics in medicine 39, 2815–2842 (2020).

30 Vink, G., Frank, L. E., Pannekoek, J. & Van Buuren, S. Predictive mean matching imputation of semicontinuous variables. Statistica Neerlandica 68, 61–90 (2014).

31 van Dijk, B. C. et al. Relationship between the Asthma Control Test (ACT) and other outcomes: a targeted literature review. BMC pulmonary medicine 20, 1–9 (2020).

32 Schatz, M. & Camargo Jr, C. A. The relationship of sex to asthma prevalence, health care utilization, and medications in a large managed care organization. Annals of allergy, asthma & immunology 91, 553–558 (2003).

