## Supplemental material for "Randomized attention-placebo controlled trial of a digital self-management platform for adult asthma"

**Supplement table 1. Baseline characteristics of individuals in modified intention-to-treat analysis**

|  | Intervention  (n=133) | Control  (n=129) | All  (n=262) |
| --- | --- | --- | --- |
| Age | 34.14 (11.82) | 35.55 (13.54) | 34.84 (12.69) |
| Gender |  |  |  |
| Female | 104 (78.20) | 101 (78.29) | 205 (78.24) |
| Male | 23 (17.29) | 26 (20.16) | 49 (18.70) |
| Other | 6 (4.51) | 2 (1.55) | 8 (3.05) |
| Depression duration |  |  |  |
| <1 month | 2 (1.50) | 1 (0.78) | 3 (1.15) |
| 1 to <3 months | 4 (3.01) | 4 (3.10) | 8 (3.05) |
| 3 months to <1 year | 9 (6.77) | 8 (6.20) | 17 (6.49) |
| 1 year to <2 years | 9 (6.77) | 7 (5.43) | 16 (6.11) |
| 2 years to <5 years | 20 (15.04) | 11 (8.53) | 31 (11.83) |
| >5 years | 89 (66.92) | 98 (75.97) | 187 (71.37) |
| Physician contact |  |  |  |
| Regular | 43.(32.33) | 41 (31.78) | 84 (32.06) |
| Occasional | 76 (57.14) | 70 (54.26) | 146 (55.73) |
| Not anymore | 12 (9.02) | 12 (9.30) | 24 (9.16) |
| Never | 2 (1.50) | 6 (4.65) | 8 (3.05) |
| Diagnosed by a physician |  |  |  |
| Yes | 131 (98.50) | 124 (96.12) | 255 (97.33) |
| No | 2 (1.50) | 5 (3.88) | 7 (2.67) |
| ACT total score^1^ | 13.01 (4.24) | 12.97 (4.42) | 12.99 (4.32) |
| SF-12 physical health subscale^2^ | 39.94 (8.85) | 40.02 (8.53) | 39.98 (8.67) |
| SF-12 mental health subscale^3^ | 38.29 (9.49) | 38.50 (10.53) | 38.39 (9.99) |

Data are n (%) or mean (SD). ^1^Patient Health Questionnaire, 8-item version (possible range 0–24), ^2^Short-Form Health Survey-12 physical health subscale (possible range 0–100), ^3^Short-Form Health Survey-12 mental health subscale (possible range 0–100).

**Supplement table 2. Outcomes**

|  | **Effect estimate** | **p-value** |
| --- | --- | --- |
| **Per protocol (N=152)** |  |  |
| ACT at 8 weeks^1^ | 1.91 (0.31 to 3.51) | 0.020 |
| Remission at 8 weeks^2^ | 1.47 (0.75 to 2.90) | 0.264 |
| ACT repeated measures (2, 4, 6, 8 weeks)^1^ | 1.34 (0.15 to 2.53) | 0.028 |
| Remission repeated measures (2, 4, 6, 8 weeks)^2^ | 1.67 (0.62 to 4.52) | 0.311 |
| SF-12 physical component score at 8 weeks^1^ | 0.81 (-1.49 to 3.10) | 0.488 |
| SF-12 mental component score at 8 weeks^1^ | 0.84 (-1.97 to 3.65) | 0.557 |
| SF-12 physical component score repeated measures (4, 8 weeks)^1^ | 0.611 (-1.32 to 2.54) | 0.534 |
| SF-12 mental component score repeated measures (4, 8 weeks)^1^ | 0.91 (-1.21 to 3.03) | 0.399 |
| MCID at 8 weeks^2^ | 2.38 (1.20 to 4.70) | 0.013 |
| Worse at 8 weeks^2^ | 0.55 (0.23 to 1.32) | 0.182 |
| **Intention to treat, multiple imputation for missing outcomes (N=262)** |  |  |
| ACT at 8 weeks^1^ | 1.56 (0.32 to 2.79) | 0.013 |
| Remission at 8 weeks^2^ | 1.45 (0.80 to 2.63) | 0.215 |
| ACT repeated measures (2, 4, 6, 8 weeks)^1^ | 1.23 (0.33 to 2.12) | 0.007 |
| Remission repeated measures (2, 4, 6, 8 weeks)^2^ | 1.40 (0.85 to 2.32) | 0.187 |
| SF-12 physical component score at 8 weeks^1^ | 0.58 (-1.45 to 2.60) | 0.578 |
| SF-12 mental component score at 8 weeks^1^ | 0.73 (-1.75 to 3.22) | 0.564 |
| SF-12 physical component score repeated measures (4, 8 weeks)^1^ | 0.75 (-1.15 to 2.64) | 0.440 |
| SF-12 mental component score repeated measures (4, 8 weeks)^1^ | 0.23 (-1.93 to 2.40) | 0.833 |
| MCID at 8 weeks^2^ | 2.17 (1.25 to 3.78) | 0.006 |
| Worse at 8 weeks^2^ | 0.76 (0.39 to 1.56) | 0.451 |
| **Intention to treat, last observation carried forward for missing outcomes (N=262)** |  |  |
| ACT at 8 weeks^1^ | 1.17 (0.02 to 2.31) | 0.046 |
| Remission at 8 weeks^2^ | 0.92 (0.53 to 1.57) | 0.753 |
| ACT repeated measures (2, 4, 6, 8 weeks)^1^ | 1.03 (0.12 to 1.93) | 0.027 |
| Remission repeated measures (2, 4, 6, 8 weeks)^2^ | 0.88 (0.41 to 1.93) | 0.762 |
| SF-12 physical component score at 8 weeks^1^ | 0.17 (-1.67 to 2.02) | 0.853 |
| SF-12 mental component score at 8 weeks^1^ | 0.61 (-1.63 to 2.85) | 0.592 |
| SF-12 physical component score repeated measures (4, 8 weeks)^1^ | -0.13 (-2.21 to 1.95) | 0.904 |
| SF-12 mental component score repeated measures (4, 8 weeks)^1^ | 0.76 (-1.69 to 3.21) | 0.543 |
| MCID at 8 weeks^2^ | 1.95 (1.17 to 3.24) | 0.010 |
| Worse at 8 weeks^2^ | 0.65 (0.35 to 1.21) | 0.172 |

^1^β-coefficient, ^2^odds ratio
